# Impact of Vaccination, Prior Infection, and Therapy on Delta and Omicron Variants

**DOI:** 10.1101/2022.03.24.22272901

**Authors:** Xiaofeng Wang, Joe Zein, Xinge Ji, Dan-Yu Lin

**Affiliations:** Lerner Research Institute and Cleveland Clinic, Cleveland, OH; Respiratory Institute, Cleveland Clinic, Cleveland, OH; Gillings School of Global Public Health, University of North Carolina, Chapel Hill, NC

## Abstract

We studied 249,070 patients who were tested for SARS-CoV-2 in the Cleveland Clinic Health System between October 1, 2021 and January 31, 2022. We found that vaccination, especially with recent boosting, was more effective than prior infection and monoclonal antibody therapy against both the delta and omicron variants. Vaccination and prior infection were much less effective against infection with the omicron variant than with the delta variant, but the opposite was true of death after infection. Boosting greatly increased the effectiveness of the two mRNA vaccines against both infection and death, although its effects waned markedly after 6 months. In addition, monoclonal antibody therapy was notably less effective at preventing death from the omicron variant than from the delta variant. Finally, the relatively low mortality of the omicron variant was due to both the reduced lethality of this variant and the increased population immunity acquired from booster vaccination and previous infection.

---

The B.1.1.529 (omicron) variant of severe acute respiratory syndrome coronavirus 2 (SARS-CoV-2), first detected in South Africa on November 25, 2021, has spread rapidly around the world and has been the predominant strain in the United States since mid-December 2021.^1^ The omicron variant contains multiple mutations that are associated with higher transmissibility and greater potential to evade immunity from neutralizing antibodies, as compared to the B.1.617.2 (delta) variant.^2-3^ We recently conducted a large study in the United States to compare the effects of Covid-19 vaccination and prior SARS-CoV-2 infection on infection with the omicron versus delta variants, and also to compare the effects of vaccination, prior infection, and monoclonal antibody therapy on death caused by the two variants.

We considered all patients who were tested for SARS-CoV-2 by polymerase chain reaction in the Cleveland Clinic Health System between October 1, 2021 and January 31, 2022. We extracted each patient’s medical record from the Covid-19 registry database.^4^ We divided the study period into two: October 1–December 11 (the delta-predominant period) and December 19–January 31 (the omicron-predominant period), omitting the week ending on December 18, when the omicron variant was emerging. A total of 133,766 and 115,304 patients were tested, with 27,001 and 45,233 positive results, during the delta- and omicron-predominant periods, respectively. The patients’ demographic and clinical characteristics are shown in Table 1.

**Table 1.** Demographic and Clinical Characteristics of the Patients Who Were Tested for SARS-CoV-2 in the Cleveland Clinic Health System During the Delta- and Omicron-Predominant Periods. The data are represented as number (%).

| Variable | Total | Dose 2 |  | Dose 3 |  | Other | SARS-CoV-2 | Death |
| --- | --- | --- | --- | --- | --- | --- | --- | --- |
|  |  | < 180 days | ≥ 180 days | < 180 days | ≥ 180 days | Vaccination |  |  |
|  | 249,070 | 22,885 (9.2) | 63,249 (25.4) | 42,637 (17.0) | 4,840 (1.8) | 15,403 (6.2) | 72,224 (29.0) | 1,088 (1.5) |
| Time period |  |  |  |  |  |  |  |  |
| Delta | 133,766 (54) | 15,028 (66) | 35,931 (57) | 11,170 (26) | 2,390 (49) | 8,049 (52) | 27,001 (37.4) | 631 (58.0) |
| Omicron | 115,304 (46) | 7,857 (34) | 27,318 (43) | 31,467 (74) | 2,450 (51) | 7,354 (48) | 45,223 (62.6) | 457 (42.0) |
| Location |  |  |  |  |  |  |  |  |
| Ohio | 216,434 (87) | 20,431 (89) | 55,909 (88) | 37,274 (87) | 4,068 (84) | 13,420 (87) | 65,761 (91.1) | 986 (90.6) |
| Other | 32,636 (13) | 2,454 (11) | 7,340 (12) | 5,363 (13) | 772 (16) | 1,983 (13) | 6,463 (8.9) | 102 (9.4) |
| Age |  |  |  |  |  |  |  |  |
| < 65 | 186,931 (75) | 20,015 (87) | 43,476 (69) | 22,115 (52) | 2,208 (46) | 12,187 (79) | 58,927 (81.6) | 266 (24.4) |
| ≥ 65 | 62,139 (25) | 2,870 (13) | 19,773 (31) | 20,522 (48) | 2,632 (54) | 3,216 (21) | 13,297 (18.4) | 822 (75.6) |
| Sex |  |  |  |  |  |  |  |  |
| Female | 144,700 (58) | 14,121 (62) | 38,738 (61) | 25,545 (60) | 2,781 (57) | 8,850 (57) | 42,345 (58.6) | 491 (45.1) |
| Male | 104,370 (42) | 8,764 (38) | 24,511 (39) | 17,092 (40) | 2,059 (43) | 6,553 (43) | 29,879 (41.4) | 597 (54.9) |
| Race and ethnicity |  |  |  |  |  |  |  |  |
| White, non-Hispanic | 169,197 (68) | 14,888 (65) | 47,128 (75) | 33,804 (79) | 4,006 (83) | 10,223 (66) | 47,921 (66.4) | 763 (70.1) |
| Black, non-Hispanic | 37,014 (15) | 4,156 (18) | 7,248 (11) | 3,785 (8.9) | 377 (7.8) | 2,774 (18) | 11,887 (16.5) | 221 (20.3) |
| Hispanic | 15,203 (6.1) | 1,907 (8.3) | 3,597 (5.7) | 1,795 (4.2) | 173 (3.6) | 1,036 (6.7) | 4,357 (6.0) | 40 (3.7) |
| Other/Unknown | 27,656 (11) | 1,934 (8.5) | 5,276 (8.3) | 3,253 (7.6) | 284 (5.9) | 1,370 (8.9) | 8,059 (11.2) | 64 (5.9) |
| Prior Infection |  |  |  |  |  |  |  |  |
| No | 231,066 (92.8) | 20,695 (90.5) | 58,578 (92.6) | 39,981 (93.8) | 4,544 (93.9) | 13,962 (91) | 68,542 (94.9) | 1,058 (97.2) |
| Yes | 18,004 (7.2) | 2,160 (9.5) | 4,671 (7.4) | 2,656 (6.2) | 296 (6.1) | 1,441 (9) | 3,682 (5.1) | 30 (2.8) |
| Smoking |  |  |  |  |  |  |  |  |
| Never | 116,546 (47) | 11,717 (51) | 31,138 (49) | 21,071 (49) | 2,323 (48) | 6,963 (45) | 33,009 (45.7) | 332 (30.5) |
| Current | 24,166 (9.7) | 2,916 (13) | 5,295 (8.4) | 2,220 (5.2) | 353 (7.3) | 2,097 (14) | 6,511 (9.0) | 99 (9.1) |
| Unknown | 108,358 (44) | 8,252 (36) | 26,816 (42) | 19,346 (45) | 2,164 (45) | 6,343 (41) | 32,704 (45.3) | 657 (60.4) |
| Respiratory conditions<sup>#</sup> |  |  |  |  |  |  |  |  |
| No | 203,311 (82) | 18,062 (79) | 50,254 (79) | 32,340 (76) | 3,623 (75) | 12,234 (79) | 12,130 (16.8) | 752 (69.1) |
| Yes | 45,759 (18) | 4,823 (21) | 12,995 (21) | 10,297 (24) | 1,217 (25) | 3,169 (21) | 60,094 (83.2) | 336 (30.9) |
| Non-Respiratory conditions<sup>#</sup> |  |  |  |  |  |  |  |  |
| No | 110,906 (45) | 9,855 (43) | 20,853 (33) | 9,599 (23) | 918 (19) | 6,301 (41) | 37,489 (51.9) | 242 (22.2) |
| Yes | 138,164 (55) | 13,030 (57) | 42,396 (67) | 33,038 (77) | 3,922 (81) | 9,102 (59) | 34,735 (48.1) | 846 (77.8) |

<sup>#</sup> Respiratory conditions: chronic pulmonary disease, pulmonary circulation disease; Non-respiratory conditions: congestive heart failure, valvular disease, peripheral vascular disease, paralysis, other neurological disorders, diabetes, hypothyroidism, renal failure, liver disease, peptic ulcer disease x bleeding, acquired immune deficiency syndrome, lymphoma, metastatic cancer, solid tumor without metastasis, rheumatoid arthritis/collagen vas, coagulopathy, obesity, fluid and electrolyte disorders, chronic blood loss anemia, deficiency anemias, alcohol abuse, drug abuse, psychoses, depression. Elixhauser Comorbidity Software, Version 3.7 was used to extract the data. The definitions and ICD-10 codes can be found at: https://www.hcup-us.ahrq.gov/toolssoftware/comorbidity/comorbidity.jsp

We used the test-negative design^5^ to evaluate the impact of vaccination and prior infection on SARS-CoV-2 infection, separately for the delta- and omicron-predominant periods. We performed multivariable logistic regression of SARS-CoV-2 infection on vaccination status, prior infection, age, sex, race/ethnicity, smoking status, comorbidities, week of testing, and geographic location. We classified vaccination status into 6 categories: 14–179 days after dose 3 of mRNA vaccine: BNT162b2 or mRNA-1273; ≥180 days after dose 3 of mRNA vaccine; 14–179 days after dose 2 of mRNA vaccine; ≥180 days after dose 2 of mRNA vaccine; any other Covid-19 vaccination; and unvaccinated.

Figure 1 shows the estimated odds ratios of SARS-CoV-2 infection with vaccination status and prior infection adjusted for other risk factors. The 3-dose series was found to be much more effective than the 2-dose series against SARS-CoV-2 infection with both the delta and omicron variants. The effects of both the 2-dose and 3-dose series waned markedly over time, and both series were considerably less effective against the omicron variant than against the delta variant. These results greatly extend existing knowledge about vaccine effectiveness.^6^ Taking the reciprocal of the odds ratio in Figure 1, we found that having dose 3 less than 180 days ago lowered the risk of infection with the delta variant (compared to the unvaccinated) by a factor of 11.1 (95% CI, 10 to 12.5), whereas having dose 2 more than 180 days ago provided little protection against infection with the omicron variant, because the omicron variant spike protein can almost completely escape from neutralizing antibodies elicited in recipients of only two mRNA vaccine doses.^7^ Prior infection lowered the risk of infection by a factor of 4.4 (95% CI: 4 to 4.8) and 1.6 (95% CI: 1.5 to 1.7) for the delta and omicron variants, respectively.

**Figure 1.**
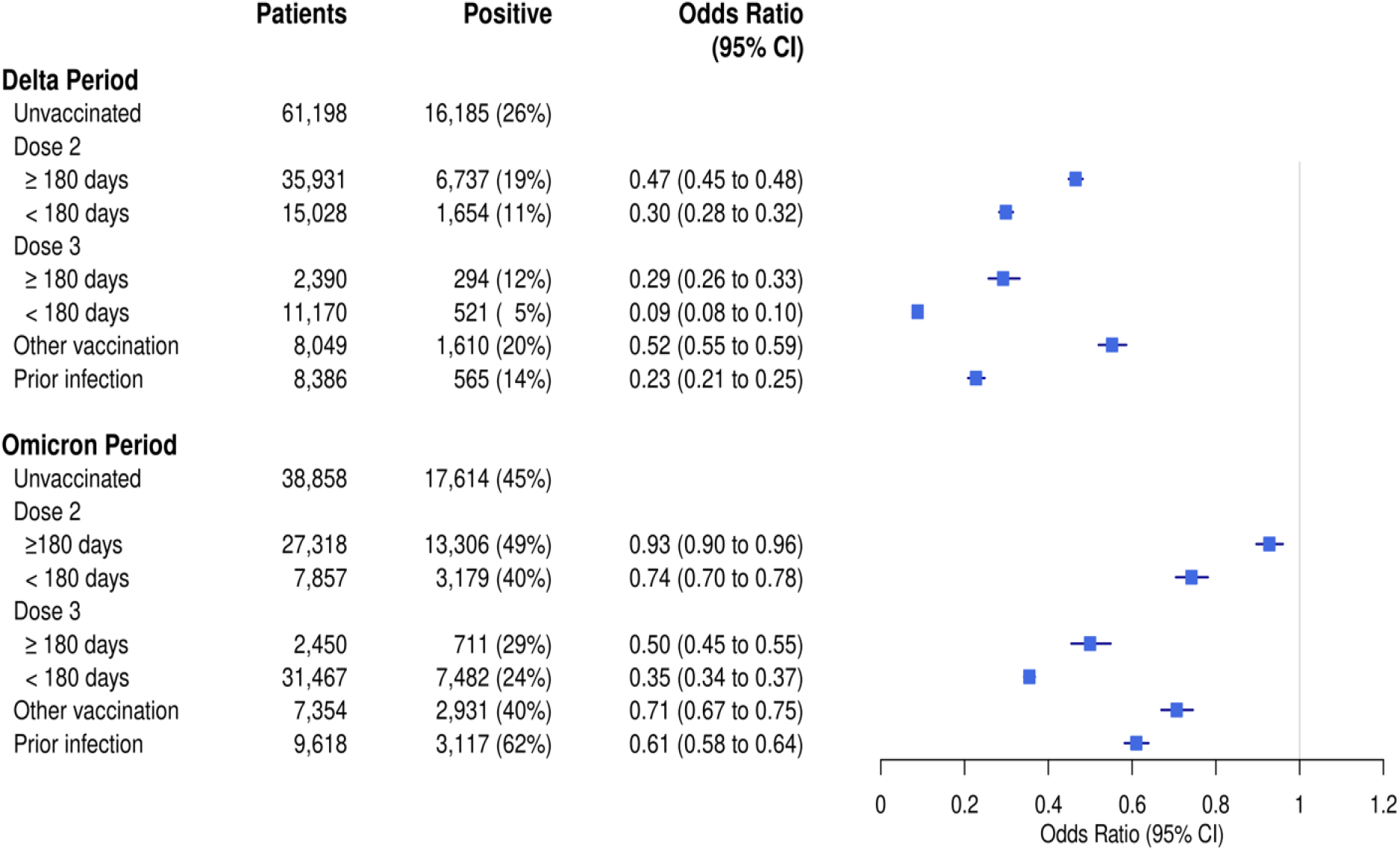
Effects of Covid-19 Vaccination and Prior SARS-CoV-2 Infection on the Risk of Infection With the Delta and Omicron Variants. The odds ratio estimates are shown in squares, and the 95% confidence intervals (95% CI) are shown by horizontal lines.

As of February 23, 2022, a total of 631 and 457 patients had died out of those who had tested positive for SARS-CoV-2 during the delta- and omicron-predominant periods, respectively (Table 1). The Kaplan-Meier estimates of the survival probabilities among the patients who had tested positive for SARS-CoV-2 infection during the omicron-predominant period were much higher than those of the delta-predominant period, with the 28-day mortality being 0.75% and 1.57%, respectively, and with the hazard ratio of dying from the omicron versus delta variants being 0.56 (95% CI: 0.49 to 0.63) (Figure 2A). To account for the fact that the rates of vaccination and prior infection were higher during the omicron-predominant period than during the delta-predominant period, we further considered the subset of patients who had not been vaccinated or previously infected. Then the survival curves between the two time periods were much closer to each other, and the hazard ratio became 0.71 (95% CI: 0.60 to 0.85) (Figure 2B), indicating that the omicron variant is intrinsically less lethal than the delta variant.

**Figure 2.**
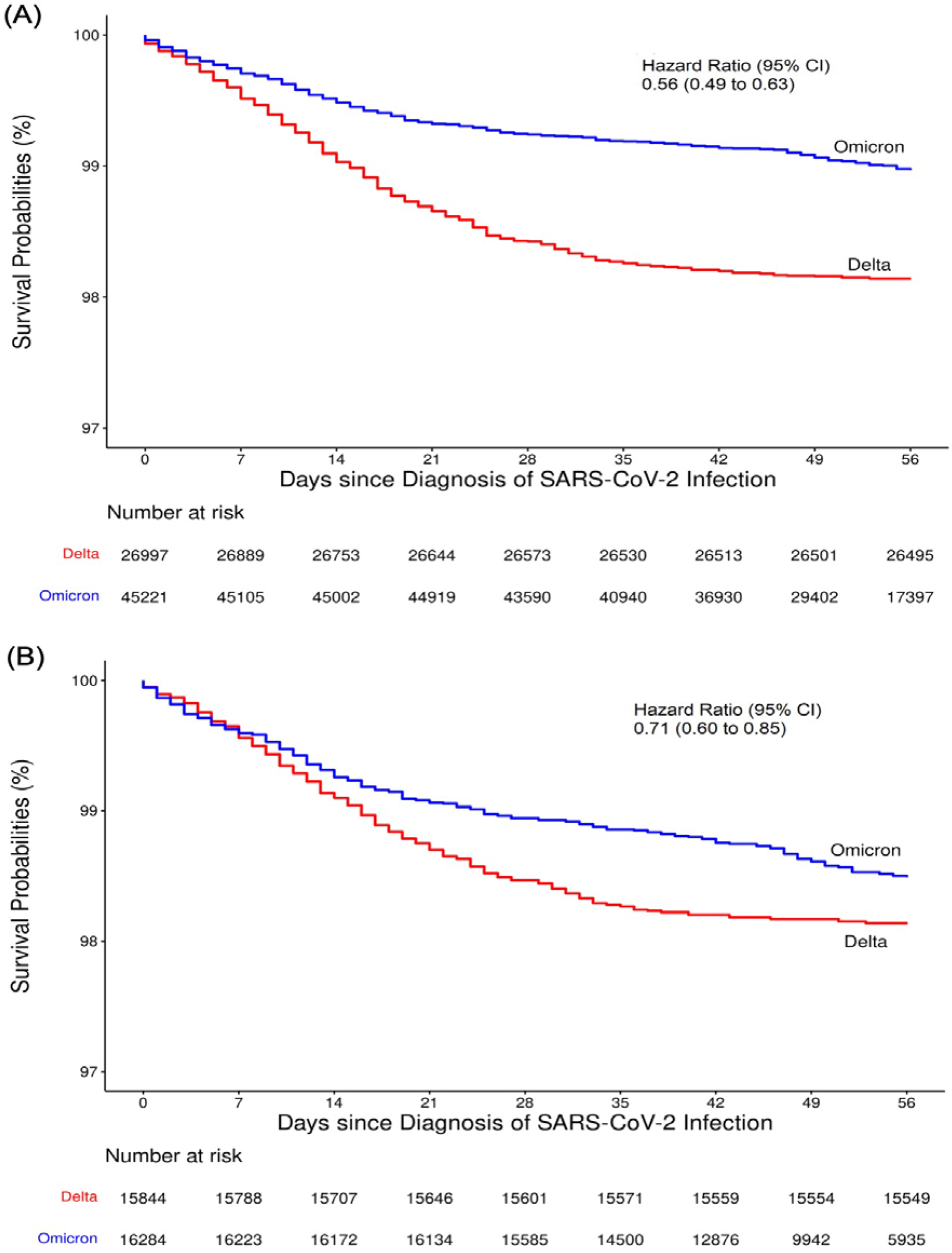
Kaplan-Meier Estimates of the Survival Probabilities for Patients Who Had Tested Positive for SARS-CoV-2 During the Delta- and Omicron-Predominant Periods: (A) All patients; (B) those not vaccinated or previously infected.

To compare the effects of vaccination status, prior SARS-CoV-2 infection and monoclonal antibody therapy on death from the omicron versus delta variants, we used multivariable Cox regression to relate the risk of death to those three variables and other risk factors separately for the delta- and omicron-predominant periods. Table 2 shows the estimated hazard ratios of death with vaccination status, prior SARS-CoV-2 infection, monoclonal antibody therapy, and other risk factors, separately for the omicron and delta variants. Older adults, males, smokers, and patients with comorbidities were at elevated risk of death from both variants. The effects of the two mRNA vaccines on death, especially for the 3-dose series, decreased over time. The 3-dose series reduced the risk of death from the omicron variant much more than from the delta variant. Taking the reciprocal of the hazard ratio in Table 2, we found that having dose 3 less than 180 days ago lowered the risk of death from the delta and omicron variants by a factor of 4.2 (95% CI: 1.9 to 9.3) and 6.7 (95% CI: 2.5 to 17.9), respectively; and having dose 3 more than 180 days ago had only a moderate effect on the risk of death from the delta variant, partly because many of those patients were immuno-compromised. Prior infection reduced the risk of death from the omicron variant by a factor of 2.9 (95% CI: 1.4 to 5.9). By contrast, prior infection had virtually no effect on the risk of death from the delta variant, because it greatly reduced the risk of infection with the delta variant, such that the patients who still got infected might have had weakened immune systems. Monoclonal antibody treatments lowered the risk of death by a factor of 2.4 (95% CI: 1.7 to 3.4) and 1.9 (95% CI: 1.1 to 3.5) for the delta and omicron variants, respectively.

**Table 2.** Effects of Covid-19 Vaccination, Prior SARS-CoV-2 Infection, and Monoclonal Antibody Therapy on the Risk of Death Caused by the Delta and Omicron Variants.

| **Variable** | **Delta Variant**<br>**Hazard Ratio (95 CI)** | **Omicron Variant**<br>**Hazard Ratio (95 CI)** |
| --- | --- | --- |
| Vaccination status |  |  |
| Unvaccinated | Reference | Reference |
| Dose 2 ≥ 180 days | 0.43 (0.29 to 0.64) | 0.43 (0.25 to 0.74) |
| Dose 2 < 180 days | 0.42 (0.34 to 0.51) | 0.40 (0.32 to 0.51) |
| Dose 3 ≥ 180 days | 0.77 (0.53 to 1.13) | 0.23 (0.17 to 0.31) |
| Dose 3 < 180 days | 0.24 (0.11 to 0.54) | 0.15 (0.06 to 0.40) |
| Other vaccination | 0.87 (0.64 to 1.19) | 0.74 (0.53 to 1.04) |
| Prior infection | 1.08 (0.70 to 1.68) | 0.34 (0.17 to 0.69) |
| Monoclonal antibodies | 0.42 (0.29 to 0.60) | 0.52 (0.29 to 0.95) |
| Age ≥ 65 (vs. < 65) | 10.1 (8.32 to 12.2) | 18.6 (14.5 to 23.9) |
| Male (vs. Female) | 1.49 (1.27 to 1.74) | 1.58 (1.31 to 1.91) |
| Race and ethnicity |  |  |
| White, non-Hispanic | Reference | Reference |
| Black, non-Hispanic | 1.19 (0.97 to 1.47) | 1.31 (1.05 to 1.64) |
| Hispanic | 0.95 (0.60 to 1.49) | 0.94 (0.59 to 1.50) |
| Other/Unknown | 0.81 (0.57 to 1.15) | 0.69 (0.46 to 1.03) |
| Smoking status |  |  |
| Never | Reference | Reference |
| Current | 1.26 (0.94 to 1.71) | 1.30 (0.91 to 1.85) |
| Unknown | 1.61 (1.35 to 1.92) | 1.55 (1.25 to 1.92) |
| Respiratory conditions | 1.37 (1.14 to 1.64) | 1.61 (1.31 to 1.99) |
| Non-respiratory conditions | 2.13 (1.73 to 2.62) | 1.75 (1.36 to 2.26) |
| Other location (vs. Ohio) | 1.23 (0.87 to 1.74) | 1.18 (0.89 to 1.57) |

By combining the results in Figure 1 and Table 2, we can estimate the overall effectiveness of vaccination and prior infection in reducing the risk of acquiring Covid-19 and then dying from it. Specifically, receiving dose 3 less than 180 days ago lowered this risk (compared to the unvaccinated) by a factor of 46.3 and 19.2 for the delta and omicron variants, respectively, and having a prior infection reduced this risk by a factor of 4.1 and 4.8 for the delta and omicron variants, respectively.

In summary, vaccination, especially with recent boosting, was more effective than prior infection and monoclonal antibody therapy against both the omicron and delta variants. Vaccination and prior infection were considerably less effective against infection with the omicron variant than with the delta variant, but the opposite was true of death after infection. Boosting greatly increased the effectiveness of the two mRNA vaccines against both infection and death, although its effects waned markedly over time. In addition, monoclonal antibody therapy was appreciably less effective at preventing death from the omicron variant than from the delta variant. Finally, the relatively low mortality caused by the omicron variant, as compared to the delta variant, was due to both the reduced lethality of the omicron variant and the increased population immunity acquired from booster vaccination and previous infection.

## Data Availability

All data produced in the present study are available upon reasonable request to the authors

